# Ceftriaxone-resistant *Neisseria gonorrhoeae* detected in England, 2015 to 2024; an observational study

**DOI:** 10.1101/2024.08.12.24311674

**Authors:** Helen Fifer, Michel Doumith, Luciana Rubinstein, Laura Mitchell, Mark Wallis, Selena Singh, Gurmit Jagjit Singh, Michael Rayment, John Evans-Jones, Alison Blume, Olamide Dosekun, Kenny Poon, Achyuta Nori, Michaela Day, Rachel Pitt, Suzy Sun, Prarthana Narayanan, Emma Callan, Anna Vickers, Jack Minshull, Kirsty Bennet, James E.C. Johnson, John Saunders, Sarah Alexander, Hamish Mohammed, Neil Woodford, Katy Sinka, Michelle Cole

**Author notes:** Corresponding author Helen Fifer, Address: UK Health Security Agency, 61 Colindale Avenue, London NW9 5EQ.

## Abstract

**Background:** Since June 2022, there has been a rise in the number of ceftriaxone resistant *Neisseria gonorrhoeae* cases detected in England (n = 15), of which one third were extensively-drug resistant (XDR). We describe the demographic and clinical details of the recent cases and investigate the phenotypic and molecular characteristics of the isolates. For a comprehensive overview, we also reviewed 16 ceftriaxone-resistant cases previously identified in England since December 2015 and performed a global genomic comparison of all publicly available ceftriaxone-resistant *N. gonorrhoeae* strains with mosaic *penA* alleles.

**Methods:** All *N. gonorrhoeae*isolates resistant to ceftriaxone (MIC >0.125 mg/L) were whole-genome sequenced and compared with 142 global sequences of ceftriaxone-resistant *N. gonorrhoeae*. Demographic, behavioural, and clinical data were collected, including treatment and outcomes.

**Results:** All cases were heterosexuals, and most infections were associated with travel to or from the Asia-Pacific region. However, some had not travelled outside England within the previous few months. There were no ceftriaxone genital treatment failures, but 3/5 pharyngeal infections and the only rectal infection failed treatment. The isolates represented 13 different multi-locus sequence types (MLSTs), and most had the mosaic *penA*-60.001 allele. The global genomes clustered into eight major phylogroups, with regional associations. All XDR isolates belonged to the same phylogroup, represented by MLST 16406.

**Conclusion:** Ceftriaxone resistance in *N. gonorrhoeae* continues to be associated with the *penA*-60.001 allele within multiple genetic backgrounds and with widespread dissemination in the Asia-Pacific region. Heightened surveillance activities have been initiated to detect further cases with the aim of interrupting further transmission.

## Introduction

Gonorrhoea is the second most common sexually transmitted infection (STI) in England, with over 85,000 diagnoses in 2023.^1^ Untreated gonorrhoea can lead to pelvic inflammatory disease, infertility and ectopic pregnancy, and can increase the risk of HIV transmission. *Neisseria gonorrhoeae*, the causative pathogen of gonorrhoea, has developed resistance to successive classes of antibiotics, and few antimicrobials remain effective in its treatment.^2,3^ Extended-spectrum cephalosporins, such as ceftriaxone, are the last-line option for empirical monotherapy.^2^ The first ceftriaxone resistant gonococcal strain, H041, was detected in Japan in 2009^4^ but failed to disseminate further. Four more ceftriaxone-resistant *N. gonorrhoeae* presented a similar picture from 2010 to 2014;^5–8^ strains were detected but failed to disseminate. The exception was strain F89, which was first isolated in France during 2010^5^ and was subsequently isolated in Spain in 2011,^6^ albeit with undetected cases along transmission chains. Unfortunately, the sporadic nature of ceftriaxone-resistant *N. gonorrhoeae* was relatively short-lived, with the emergence of the FC428 strain in Japan during 2015.^9^ The FC428 strain is associated with ceftriaxone resistance determined by the mosaic *penA*-60.001 allele that encodes the gonococcal penicillin-binding protein 2. Strains that cluster within the FC428 clone have been detected in numerous countries in the Asia-Pacific region including China, Cambodia, and Vietnam, as well as in Canada and across Europe,^10,11^ with the first case detected in the United Kingdom (UK) in 2018.^12^ The *penA*-60.001 allele has also been detected in different genomic backbones,^10,11,13^ suggesting independent recombination events with a *penA*-60.001 donor, such as commensal *N. subflava*,^14^ or recombination with other FC428-like isolates. The first ceftriaxone-resistant *N. gonorrhoeae* strain with high-level resistance to azithromycin (MIC ≥256 mg/L), considered to be extensively-drug resistant (XDR), was isolated during 2018 in both the UK and Australia and harboured the *penA*-60.001 allele, but was unrelated to the FC428 clone.^13,15^ There were no identified links between the individuals, thus highlighting the extent of antimicrobial resistance (AMR) surveillance gaps.^15^

Previously we reported a concerning increase in ceftriaxone-resistant *N. gonorrhoeae* cases in the UK, with 10 cases detected in the six-month period between December 2021 and June 2022, compared with nine cases detected between December 2015 and September 2021.^16^ Most of these cases were associated with travel to or from the Asia-Pacific region, and all were heterosexual individuals. Alarmingly, a further 15 cases have been detected in England between June 2022 and May 2024, including five cases which were XDR, marking the highest number of XDR cases reported in any European country to date. Here we describe the demographic and clinical details of these most recent cases, and phenotypic and molecular characteristics of the isolates. For a comprehensive overview, we have also included details for 16 ceftriaxone-resistant cases previously identified in England, 12 of which have been published,^12,13,16,17^ and performed a global genomic comparison of all publicly available ceftriaxone-resistant *N. gonorrhoeae*strains with mosaic *penA* alleles.

## Methods

### Data collection

Data were collected as part of case management and outbreak control, and included sex, age, sexual orientation, travel history, site of infection, symptoms, treatment, test of cure (TOC), number of partners, and partner outcome. Data collection was done by the clinical team in the sexual health service (SHS) and was shared securely with the UK Health Security Agency (UKHSA) to support clinical and public health management.

### Ethics

The Research Ethics and Governance Group of the UK Health Security Agency waived ethical approval for this work. This analysis was undertaken for health protection purposes under permissions granted to UKHSA to collect and process confidential patient data under Regulation 3 of The Health Service (Control of Patient Information) Regulations 2020 and Section 251 of the National Health Service Act 2006.

### N. gonorrhoeae isolates

The UK gonorrhoea management guideline recommends that specimens are taken for culture from all individuals diagnosed with gonococcal infection, prior to treatment.^18^ Primary diagnostic laboratories in England are requested to send all *N. gonorrhoeae* isolates found to be resistant to ceftriaxone (MIC >0.125 mg/L) to the UKHSA STI Reference laboratory (STIRL). Isolates received at STIRL were confirmed as *N. gonorrhoeae* by MALDI-TOF (Bruker, Coventry, UK) and MICs were determined using Etest (bioMerieux, Basingstoke, UK) on GC agar (BD, Wokingham, UK) supplemented with 1% Vitox (Oxoid, Basingstoke, UK). MICs were interpreted with EUCAST clinical breakpoints, where available.^19^ If an isolate was found to be ceftriaxone resistant at the primary laboratory but was not retrievable to send to STIRL, residual nucleic acid amplification test (NAAT) specimens were sent to STIRL, when available, for *penA*-60.001 PCR.^20^ All cases of ceftriaxone resistance confirmed by either culture (n=29) or PCR (n=2) at STIRL were included in this study.

### Whole-genome sequencing

Genome sequences of all ceftriaxone-resistant isolates in England confirmed by STIRL between January 2015 and May 2024 (n = 29) including those published previously (n = 12/29) were included in this study. Two cases were confirmed only by *penA*-60.001 PCR and an isolate was not available for sequencing.

Genomic DNA of the 13 recent isolates (June 2022 - May 2024), as well as 4 isolates detected between 2017 and 2021 that had not previously been sequenced, was extracted using the Qiasymphony SP using the DSP DNA mini kit (Qiagen, Manchester, UK) and subsequently genome sequenced on Illumina platforms (Cambridge, UK) using the paired-end protocol. To put these isolates into a global context, short read datasets (n = 116) and assembled genomes (n = 26) of publicly available ceftriaxone-resistant isolates with mosaic *penA* alleles were downloaded from the European Nucleotide Archive (ENA) and National Center for Biotechnology Information (NCBI) databases. Retrieved genome sequences were from ceftriaxone-resistant isolates identified between 2009 and 2024 in 16 countries (Supplementary data, Table S1). The full-length alignment of reads and assembled genomes against the reference genome FA1090 (GenBank AE004969) was generated using Snippy (**GitHub - tseemann/snippy**) and then used as an input for Gubbins (https://github.com/nickjcroucher/gubbins) to remove recombination. The maximum likelihood tree was generated from the core filtered alignment generated by Gubbins using RAxML. Sequencing reads from STIRL isolates were also assembled using SPAdes (https://github.com/ablab/spades) and the generated contigs were scanned against the multilocus sequence typing (MLST, https://pubmlst.org), *N. gonorrhoeae* multiantigen sequence typing (NG-MAST, https://pubmlst.org) and antimicrobial resistance (NG-STAR, https://ngstar.canada.ca) typing schemes using BlastN. Known antimicrobial resistance determinants were also checked by BlastN using the *N. gonorrhoeae* reference database of Pathogenwatch (**GitHub - pathogenwatch/amr-libraries**). Mutations in 23S rRNA and number of mutated copies were checked by mapping using gene finder (**GitHub – ukhsa-collaboration/gene_finder**). Sequencing reads generated in this study were submitted to the European Nucleotide Archive under project number PRJEB76977.

## Results

### Patient characteristics and treatment outcomes

Thirty-one cases of ceftriaxone-resistant gonococcal infection in England were confirmed by STIRL between 2015 and May 2024 (Table 1). All were heterosexual people living in England, mainly in their 20s, although five were over 40 years old. Of the recent 15 cases (H22-324 – H24-403), most had acquired the infection in the Asia-Pacific region. However, there were four cases who had not; one case had travelled to Bulgaria (H24-541), but the others had not travelled outside the UK within the previous few months (H24-403, H24-403A, H23-672). The 15 cases comprised three partnerships and nine individuals. Overall, there were several partners in the UK who could not be contacted (data not shown).

**Table 1.**
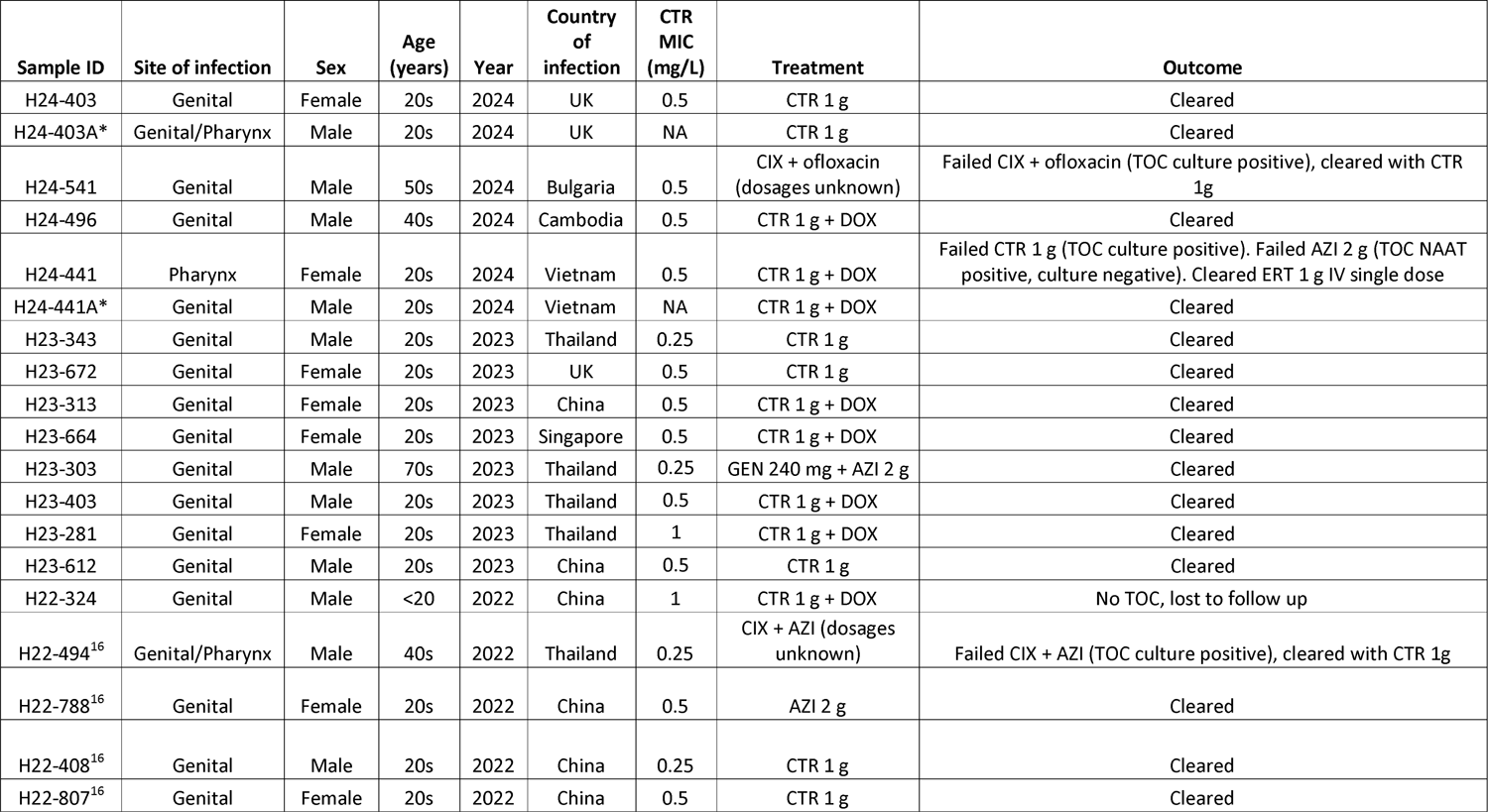
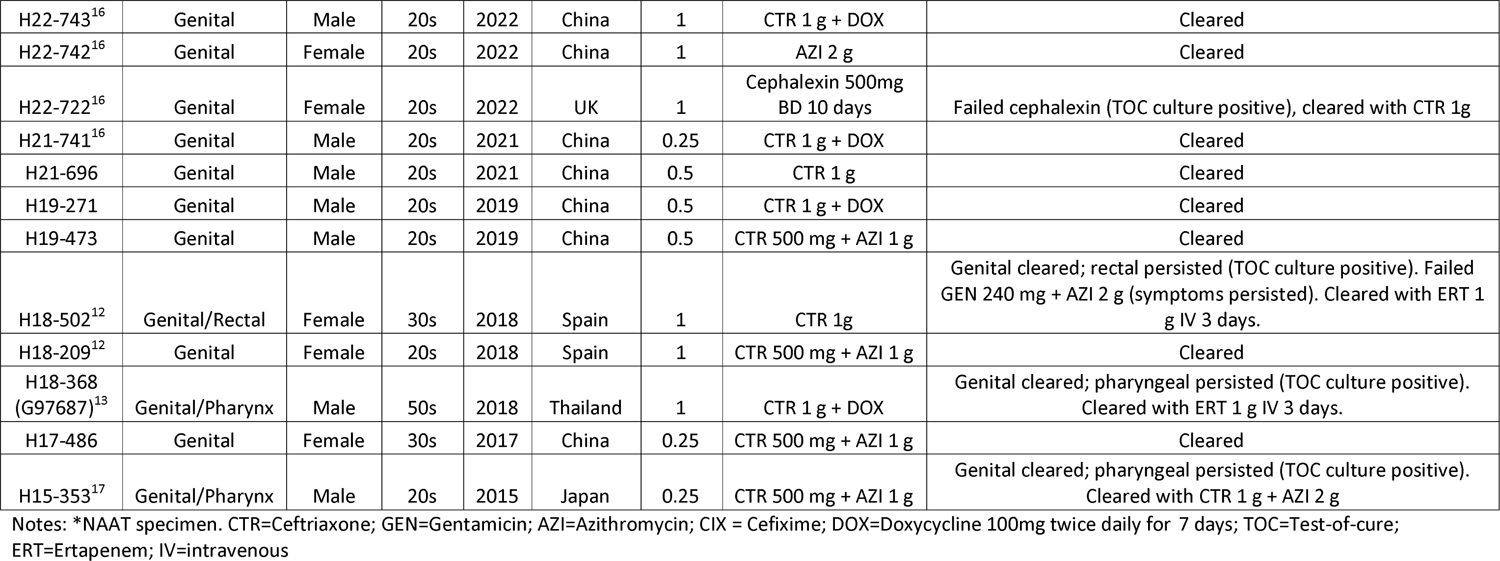
Demographic, clinical and treatment data for 31 cases of ceftriaxone resistant *N. gonorrhoeae* identified in England, 2015 to 2024.

Of the 15 most recent cases, all genital infections and one pharyngeal infection were successfully treated with ceftriaxone 1 g intramuscularly (IM). Some individuals also received doxycycline 100 mg twice daily for seven days for confirmed or presumptive chlamydia co-infection. One female case with a pharyngeal infection (H24-441; ceftriaxone MIC 0.25 mg/L, azithromycin MIC 0.125 mg/L) failed treatment with ceftriaxone 1 g IM, with a positive pharyngeal NAAT and culture three weeks later. She was then given azithromycin 2 g orally but had a positive pharyngeal NAAT four weeks after this treatment. There was no risk of reinfection. The cycle threshold (Ct) values of the NAAT were reviewed and showed little change from the first specimen (Ct 20.7 to 25.9), indicating persistent infection. She was then given a single dose of ertapenem 1 g intravenously (IV) but had positive NAATs for several weeks following this. However, the Ct values were higher (36.9 at three weeks post-ertapenem, 33.9 at five weeks) then negative at seven weeks. It was likely that the persistent low-level positivity post ertapenem was due to residual DNA. Overall, in this case series since 2015, three of five pharyngeal infections and the only rectal infection failed treatment.

### N. gonorrhoeae isolates

Twenty-nine isolates were confirmed to be ceftriaxone resistant by STIRL. There were two additional cases where the isolates had been reported as ceftriaxone resistant by the primary laboratory (and in one case also high-level azithromycin resistant) but could not be retrieved; PCR of the NAAT specimens (H24-441A and H24-403A) confirmed these isolates carried the *penA*-60.001 allele.

Moreover, the isolates from both individuals’ partners (H24-441 and H24-403) were confirmed to be ceftriaxone resistant. All ceftriaxone-resistant isolates were also resistant to cefixime and ciprofloxacin (Table 2). Nearly all were also resistant to penicillin and tetracycline. The azithromycin MICs of nine isolates were above the ECOFF (1.0 mg/L), with six isolates considered to be XDR, having high-level azithromycin resistance (MIC >256 mg/L). All isolates were susceptible to spectinomycin and had low gentamicin MICs (range 2 – 4 mg/L) and low ertapenem MICs (range 0.008 – 0.032 mg/L).

**Table 2.**
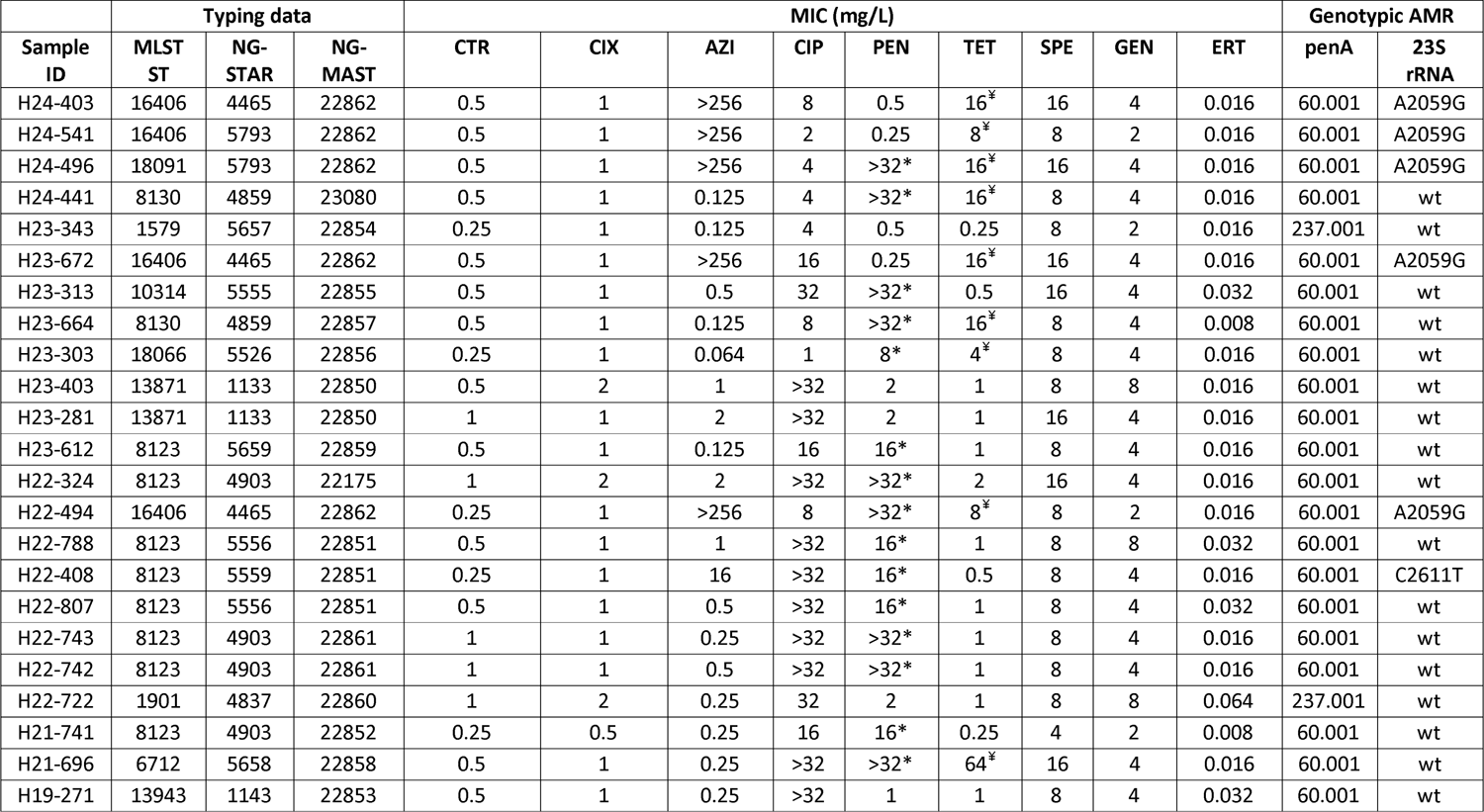
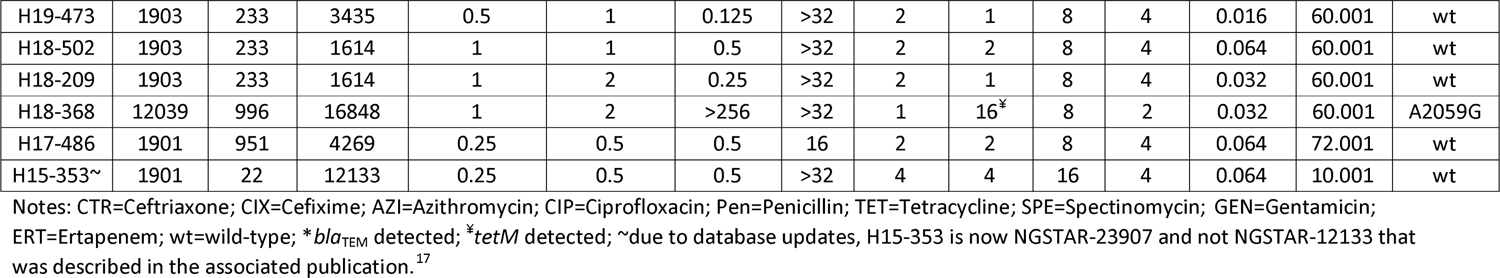
Typing data, antimicrobial susceptibility testing results and molecular determinants of resistance for 29 ceftriaxone resistant *N. gonorrhoeae* isolates identified in England, 2015 to 2024.

Genome sequences identified 13 different MLSTs, including ST8123 (n = 8), ST1901 (n = 3), ST1903 (n = 3), ST8130 (n =2), ST13871 (n =2), ST16406 (n=4), ST1579 (n=1), ST6712 (n = 1), ST10314 (n =1), ST12039 (n = 1), ST13943 (n = 1), ST18066 (n = 1) and ST18091 (n=1). These were further subdivided into 19 and 21 different NG-STAR and NG-MAST types, respectively (Table 2).

Most of the 15 recent cases harboured the mosaic *penA*-60.001 allele conferring ceftriaxone resistance, apart from one isolate (H23-343; *penA*-237.001 allele) (Table 2). Isolates with high-level azithromycin resistance had the A2059G 23S rRNA gene mutation. Ciprofloxacin resistance was conferred by GyrA-91 (S91F) and GyrA-95 (D95A/G) alterations with either the ParC-86 (D86N) or ParC-87 (S87I/N/R) modification. High levels of resistance to penicillin (MIC range 8 - ≥ 32 mg/L) and tetracycline (MIC range 4 – 64 mg/L) were linked to the acquisition of *bla*_TEM_ and *tet(M)* genes, respectively. In addition, most isolates harboured alterations in the MtrR efflux regulator, that are associated with low-level resistance to azithromycin, penicillin and tetracycline, with PorB and PonA alterations also contributing to penicillin resistance.^3^ Isolates belonging to ST8123 also had a C-104T mutation in the promoter of the NorM efflux pump, previously shown to contribute to ciprofloxacin resistance (Supplementary data, Table S2).^3^

### Phylogenetic analysis

Genomes of ceftriaxone-resistant isolates (n = 171), including those identified by STIRL, belonged to 25 different MLSTs. Of these, ST1903 was the most dominant, accounting for nearly a third of the isolates (n = 56/171), followed by ST13871 (n = 24), ST 8123 (n = 15), ST7363 (n = 12), ST8130 (n = 12), ST16406 (n = 10), ST1901 (n = 6), ST1600 (n = 7), ST7365 (n=5) and 16 other STs represented by less than four genomes each. The phylogenetic analysis clustered these isolates into eight major phylogroups (clades I to VIII) in accordance with STs (Figure 1). The largest group (clade I, Figure 1) encompassed 100 isolates from 12 different countries belonging to eight different MLSTs that were all related to the spreading international FC428 clone containing *penA*-60.001. Only six STIRL isolates (H18-209, H18-502, H19-271, H19-473, H23-281 and H23-403) clustered within this widespread international clone; the majority (n = 23) were interspersed across the other major phylogroups (Figure 1). In particular, eight isolates from England (H21-741, H22-408, H22-788, H22-324, H22742, H22-743, H22-807 and H23-612) and two from Wales (H22-631 and H22-303)^16^ associated with travel from China and belonging to ST8123 from 2021 (n = 1), 2022 (n = 8) and 2023 (n = 1), clustered together in clade-VI, which also included five isolates reported from China (CD19-81, CD20-37 and CD20-64) and the USA (LRRBGS-1327 and LRRBGS-1328). Associations of travel data with the phylogeny also showed that isolate H24-496, linked to travel to Cambodia, was closely related to two isolates detected in Cambodia (22R655567T and 22R655494S), while isolate H23-313 tightly clustered with isolate GD2021027 from China. Due to lack of representativeness in the global sequenced collection, it was not always possible to confirm links to suspected regions of infection. However, all STIRL isolates linked with travel to Thailand clustered with isolates from the Asia-Pacific region (i.e., Vietnam and Cambodia). H24-441 and H23-664 isolates clustered in clade-VII which along with clade-VIII fell into the gonococcal lineage B that historically harbours more susceptible isolates when larger genomic studies are performed.^3^

Most genomes of ceftriaxone-resistant isolates (n = 158/171, 92%) carried the *penA-*60.001 allele and only a few had other *penA* alleles. In addition to *penA-*10 (n =1), *penA-*72 (n =1) and *penA-*237 (n = 3) that were identified in STIRL isolates, a handful of genomes carried *penA*-37 (n =1), *penA-*42 (n =2), *penA-*64 (n=1), *penA-*169 (n=1), *penA-*232 (n=2) or *penA-*273 (n =1). Overall, high-level azithromycin resistance associated with the A2049G substitution in 23S rRNA was only observed in a small number of ceftriaxone-resistant isolates (i.e., XDR isolates) belonging to the same phylogroup, clade IV (Figure 1). The latter included 14 isolates belonging to ST16406 (n = 10) and its single locus variants (SLV) ST18091 (n = 1) and ST12039 (n = 3). These were reported in Austria (n = 1), Australia (n = 2), Cambodia (n = 3), France (n = 2) and England (n = 6).

## Discussion

To date, all individuals with ceftriaxone-resistant gonorrhoea detected in England were heterosexual, mostly in their 20s and usually had travel links with the Asia-Pacific region. Although not all partners could be traced, fortunately there appears to have been limited onwards transmission within England. Possibly, because most cases are travel-associated and are not part of dense sexual networks, with termination of the transmission chain. However, the frequency of case detection has increased rapidly since 2021, without any changes in clinical practice or surveillance. Most notably, there has been an alarming increase of XDR cases, with an unprecedented figure of five cases detected in a recent six-month period; November 2023 – May 2024, compared to the total of seven XDR cases detected in England since 2018.

Ceftriaxone resistance in England is still mainly conferred by the *penA*-60.001 allele, although the sequence types of isolates are diverse, and tend to cluster with isolates identified in countries of the Asia-Pacific region, consistent with the reported travel links. Ceftriaxone resistant *N. gonorrhoeae* has not yet been detected in the gay or bisexual population in England. Heightened surveillance and public awareness activities are currently underway to identify cases as soon as they emerge with the aim to interrupt the transmission of cases that can impose challenging treatment situations.

Most of the globally sequenced ceftriaxone-resistant isolates belonged to the international FC428 clone. However, the phylogenetic analysis revealed multiple other phylogroups comprising isolates bearing mosaic *penA* alleles which supports the evidence that resistance to ceftriaxone has emerged across multiple genetic backgrounds^10,11,13^ that might be even more diverse than reported in this study. Early ceftriaxone-resistant isolates (identified between 2009 and 2015), including those from England, expressed various mosaic *penA* variants,^4–8^ but these were later dominated by those carrying the *penA-*60.001 allele. More recently, ST7363 isolates carrying *penA-*232.001 (which were genetically related to H041^4^ and A8806^8^ isolates reported in Japan and Australia between 2009 and 2013) have been reported in China,^21^ but none were detected in England. In contrast, ST1901 isolates carrying *penA-*237, first reported in France in 2022,^22^ were detected in two English isolates belonging to ST1901 and its SLV ST1579 (H22-722 and H23-343). Genomic analysis showed that the XDR isolates remained limited to ST16406 and its SLVs, first reported in the UK in 2018 in a heterosexual male after sexual contact in Thailand. These findings suggest that XDR *N. gonorrhoeae* isolates might be currently linked, and possibly restricted, to the global dissemination of this clade. Although a high degree of diversity is still present within the XDR clade-IV established from our analysis.

The prevalence of *N. gonorrhoeae* with ceftriaxone resistance in China was reported to be 8.1% in 2022, with five provinces reporting >10%.^23^ The Enhanced Gonococcal Antimicrobial Surveillance Programme (EGASP) in Cambodia recently reported ceftriaxone MICs ≥0.125 mg/L in 29 isolates that harboured the *penA-*60.001 allele distributed across 9 different MLSTs detected during 2021-22.^11^ Three were ST-16406 that also displayed high-level azithromycin resistance i.e., XDR.^11^ Worryingly, the Cambodian situation continues to worsen, with 15.4% ceftriaxone resistance and 6.2% XDR detected during 2022-23.^24^

In our English case series, all genital infections were successfully treated with ceftriaxone, despite the isolates being categorised as resistant to ceftriaxone, according to EUCAST breakpoints.^19^ However, some patients received dual therapy with 1 g azithromycin, and so we cannot know for certain that these infections would have cleared with ceftriaxone alone. Our dataset is currently the most comprehensive available, with full treatment outcome and site of infection data from individuals who have been infected with ceftriaxone resistant *N. gonorrhoeae*, who were treated using recommended regimens. There are very few data on the frequency of treatment failures in regions with a high prevalence of ceftriaxone resistance. One study from China of 1686 patients with uncomplicated gonorrhoea, found that all patients were cured with ceftriaxone, even though nearly 10% had an isolate with decreased susceptibility to ceftriaxone.^25^ However, it was noted that 72.7% of patients in the study were treated with a higher than standard dosage (>1 g), and it was suggested that this may be because clinicians were concerned that Chinese manufactured ceftriaxone was less potent than drug manufactured elsewhere.^25^

It is concerning that in our study, three of five pharyngeal infections and the only rectal infection failed treatment. It may be appropriate to reconsider clinical breakpoints according to the site of infection, with a higher MIC breakpoint introduced for genital infections. There is no clinical breakpoint or ECOFF for ertapenem, but it has been shown that for isolates with raised ceftriaxone MICs, the ertapenem MIC is lower,^26^ as was the case for all 29 isolates described in this paper.

However, this is not universal, particularly in the presence of a *penA* mosaic allele.^27^ In a recent clinical trial ertapenem 1 g IM was shown to be non-inferior to ceftriaxone 1 g IM, but the isolates in this trial were susceptible to ceftriaxone.^28^ To note, in the UK ertapenem is licensed for IV but not IM use. In 2018 ertapenem 1 g IV for three days was used to treat two cases of ceftriaxone treatment failure,^13^ however the most recent treatment failure in 2024 within this current study, was successfully treated with a single dose of ertapenem 1 g IV. Single dose treatment is preferable for both provider and patient.

The UK gonorrhoea management guideline recommends universal TOC at two-to-three weeks after treatment;^18^ settings which do not perform TOC will not detect asymptomatic treatment failures. Pharyngeal infection is recognised as being harder to treat, possibly because tissue penetration of antimicrobials is limited;^2^ the UK guideline recommends that all individuals with epidemiological links to the Asia-Pacific region, or anyone with urogenital ceftriaxone-resistant *N. gonorrhoeae*, should have a pharyngeal NAAT and culture taken.^18^ As described in this study, ceftriaxone 1 g IM can be expected to clear urogenital infection, but pharyngeal infection may persist and may be a potential source for onwards transmission. Without testing the pharynx these asymptomatic infections will remain undetected. Most countries’ national guidelines do not recommend routine testing of the pharynx in heterosexual individuals; the prevalence of pharyngeal gonorrhoea in heterosexuals is largely unknown, and routes of transmission remain ill-defined. It remains unclear whether kissing is a factor in gonorrhoea transmission.^29^ In this study, the four heterosexual men with pharyngeal infection denied having oral sex. Studies have reported a high prevalence of pharyngeal gonorrhoea in heterosexual men and women who were contacts of gonorrhoea cases, and that oral sex is not associated with pharyngeal infection in heterosexual men.^30^

It is likely that the detection of these cases in England represents just the tip of the iceberg of gonococcal AMR, given the large surveillance gaps globally. England has a comprehensive surveillance system including both sentinel and real-time detection of gonococcal AMR, enabled by the capacity to culture all gonococcal infections and perform universal TOC. Given that nearly all our cases had epidemiological links to the Asia-Pacific region, and that countries in the region are reporting worryingly high levels of ceftriaxone resistance,^23,24^ it is essential that action is taken at a global level. This includes access to rapid testing, quality-assured antimicrobial susceptibility testing, robust surveillance, monitoring of treatment failures and antimicrobial stewardship to maintain the viability of the current treatment options. Ultimately, additional treatment options along with innovative control strategies are required to ensure gonorrhoea remains a treatable infection.

## Data Availability

All data produced in the present study are available upon reasonable request to the authors.

## Acknowledgments

We would like to thank all microbiology laboratories for submitting isolates to the UKHSA STIRL, as well as all laboratory staff in STIRL who processed the referred gonococcal isolates.

## Contributors

HF, MDoumith and MC prepared the manuscript, and all authors contributed to the review of the final manuscript. MC, MDay, RP, AV, JM, SA and NW managed and performed the laboratory work. MDoumith developed and performed the bioinformatic analysis. SSun, PN, EC, KB, JJ, and JS performed data collection and analysis for epidemiological information. LR, LM, MW, SSingh, GJS, MR, JEJ, AB, OD, KP and AN managed the clinical cases. HF, HM, and KS coordinated the incident response teams.

---

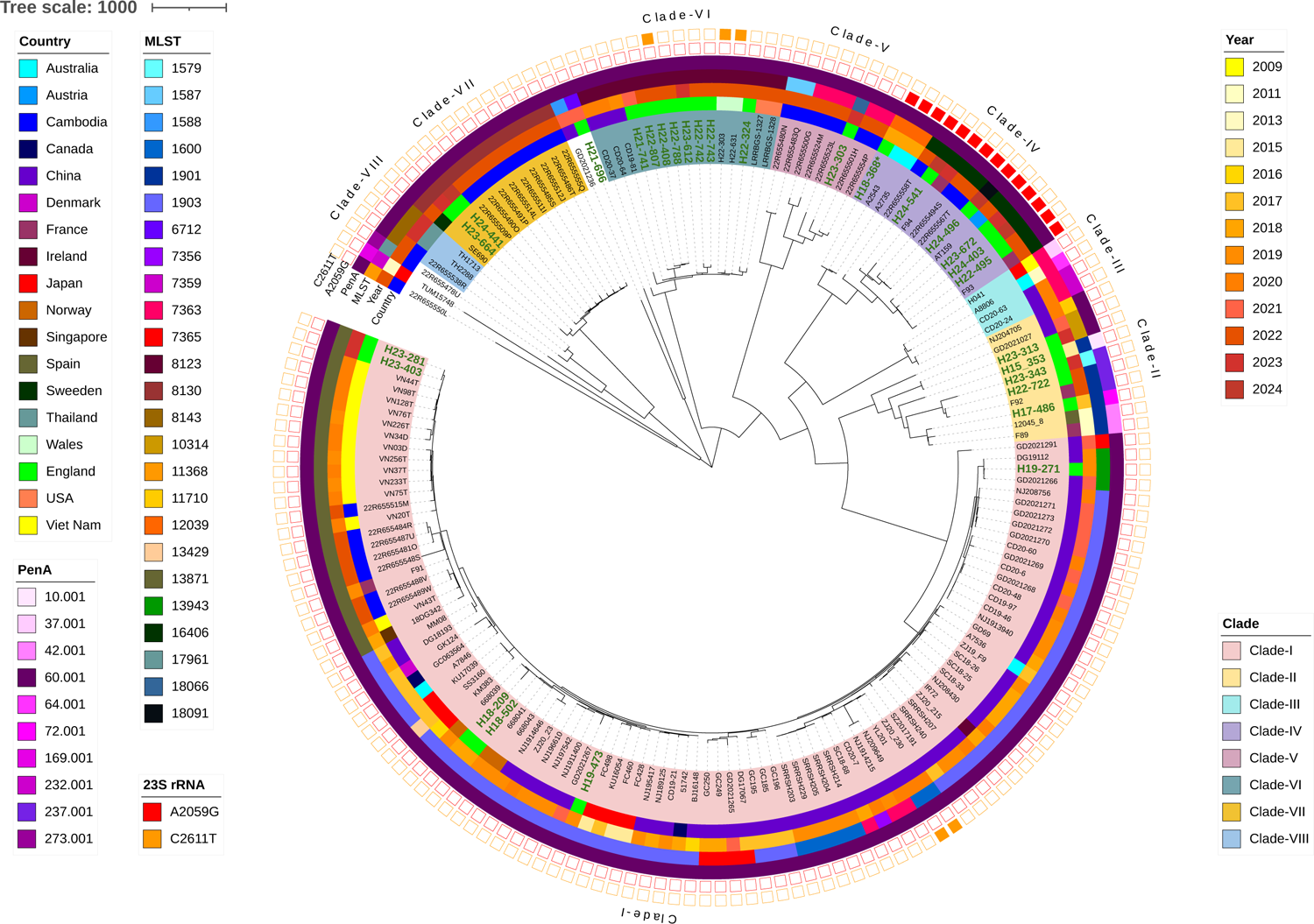

